# Assessment of outcomes in Intensive Care Unit delivery at Tibebe Ghion Specialized Hospital, North West Ethiopia

**DOI:** 10.64898/2025.12.15.25342264

**Authors:** Amanuel Sisay Endeshaw, Fantahun Tarekegn Kumie, Misganew Terefe Molla, Gashaw Abebe Zeru

**Affiliations:** Department of Anesthesia, College of Medicine and Health Science, Bahir Dar University, Ethiopia

**Keywords:** Intensive Care Unit, Mortality, Pattern, Adult

## Abstract

**Background:** The Intensive care unit (ICU) service is highly limited in sub Saharan Africa countries due to varying of quantities and qualities infrastructures. However, the burden of critical ill patient in low resource countries is higher and possibly increasing with urbanization, developing epidemics and access to hospitals service. The main objective of this study was to assess the outcomes of ICU admission and identify the areas of improvement in critical care.

**Methods:** an institutional based retrospective cross sectional study design was conducted on eligible patients who were recorded at intensive care unit registration log book from January 1, 2019 to June 30, 2020. The data was entered, edited and analyzed in stata software (version14). We performed the adjusted survival analysis between ICU mortality and independent variables by using logistic regression. We also showed patients mortality in ICU by Kaplan-Meier (KM) survival analysis.

**Results:** From January 1, 2019 to June 30, 2020 data were taken from 454 cases at Tibebe Ghion Specialized Teaching Hospital ICU registered log book through prepared questionnaire. The overall ICU mortality was 31.3%. The common leading causes of admission to ICU were head injury (19.6%) followed by non-tuberculosis respiratory problems (11.89%), post abdominal surgeries (8.37%) and myocardial infarction (6.82%). In this study, 36.3% of ICU admitted patients were served by mechanical ventilation. The logistic regression analysis ICU mortality showed that patient stayed in ICU with odds ratio (OR = 1.37 [95% CI, 1.16 – 1.62]; *P* = 0.000),needs for mechanical ventilation(OR = 0.18 [95% CI, 0.12 – 0.28]; *P* = 0.000), days on mechanical ventilation (OR = 0.73 [95% CI, 0.61 – 0.87]; *P* = 0.001) and non-infection (OR = 0.45 [95% CI, 0.24 – 0. 69]; *P* = 0.000) were associated with risk of ICU mortality.

**Conclusion:** The mortality rate of our adult ICU was 31.3% with the most common causes of admission and deaths being on head injury. The highly statistical significant predictors of ICU mortality were infection, needs for mechanical ventilation, ICU stay and days on mechanical ventilation.

## 1. Introduction

The first conception of intensive care unit has been founded by Denmark anesthetist during the polio pandemic before half of century ago(1). An intensive care unit is a separated area which is devoted to provision of care for critical illness(2). In high resources settings of the country, intensive care units (ICU) services are correlated to the better outcomes of critically ill patients. However, the high cost demanding of equipment and trained health professionals has limited the opening out of intensive care unit (ICU) in low income countries(3). In addition, there is a controversy on effectiveness of conventionally resource critical care and ICU expansion in situation where supply scarcity is faced(4).

There is an evidence of justified and appropriates of critical care in resource poor sub-Saharan Africa countries(5). Moreover, the load of critical patients in low resource countries is high and possibly rising with urbanization, evolving epidemics and access to hospitals service (6-8).Conversely, the ICU care is highly limited in sub Saharan Africa due to varying of quantities and qualities infrastructures(9, 10).

In Ethiopia, one retrospective study reported that 50.4 % of deaths was in general ICU (11), another study reported 39% in medical ICU(12), while another reported 37.7% of death in general ICU admission(13). A Tanzania study demonstrated that mortality rate of patients admitted to ICU was 41.6 % (14). A similar study of Abuja in Nigeria showed that the mortality rate of patients admitted to intensive care unit was 68.4% (15).

Progresses in critical care is depends on continuous overview of facility improvement plans with the focus of sustainability and cost effectiveness analysis (11). Therefore, research is essential in resource poor settings, like our country, Ethiopia. There have not been previous studies of mortality and morbidity conducted in Tibebe Ghion Specialized Teaching Hospital intensive care unit. The main objective of this study was to investigate the outcomes of ICU admission and to identify the areas of improvement in critical care.

## 2. Materials and methods

Tibebe Ghion Specialized Hospital is the largest new referral teaching hospital in North West Ethiopia. It started as a teaching and referral hospital on January, 2019 with 10 medical and surgical adult ICU beds and 02 separated pediatric ICU beds. This study was conducted on 10 bedded adult ICU with retrospective cross sectional study design. The data was obtained from ICU registered log book.

### Sample size and sampling methods

All sequentially admitted patients to adult ICU from January 1, 2019 to June 30, 2020 were included based on inclusion criteria of all patients who were admitted to intensive care unit with age > 12 years old at registered log book. However, Patient’s age < 12 years old, uncompleted data registration and those patients who died on arrival (2 hours of admission) were measured as exclusion criteria. The 2 hours of admission was not an adequate time to provide optimal care in ICU, and for the reason that the outcome of the patient could be related to emergency or former ward care.

### Study variables

#### Dependent variable

The outcome variable of our study was mortality in ICU services. Mortality was also designated as a binary variable (1 if died and 0 if discharged alive).

#### Independent variables

The explanatory variables of the study were age, sex, diagnosis at admission, need for mechanical ventilation, length of mechanical ventilation stay, length of stay in ICU, and diseases character (medical versus surgical and infectious versus noninfectious).

#### Data collection

The data was collected on registered log book based on: age, sex, diagnosis at admission, need for mechanical ventilation, length of mechanical ventilation stay, length of stay in ICU, and diseases character (medical versus surgical and infectious versus noninfectious). Two bachelor anesthetists were engaged for data collection and one master anesthetist as a supervisor. Training was provided for them about the objective of the study, the questionnaire’s content and confidentiality before 3 days of the review. A week before of data collection, pre-testing was done on 5% of the registered data in Tibebe Ghion Specialized Teaching Hospital ICU to check its completeness. According to the finding of the pretest, some questions were reformed to access the optimal information. Completeness of the data and its consistency were checked by the principal investigator.

### Data analysis

The data was cleaned, edited and analyzed by using stata software (version 14). The clinical characteristics and demographics were presented in the form of counts and percentages for all categorical data. Continuous variables were described as mean (SD) when normal distributed data or median (IQR) when not. We used chi square (χ^2^) and Fisher’s exact tests to compare categorical variables. Significance of the result was set at p < 0.05.

Moreover, we performed the adjusted survival analysis between ICU mortality and independent variables (age, sex, diagnosis at admission, need for mechanical ventilation, length of mechanical ventilation stay, length of stay in ICU, and diseases character) by using logistic regression. We also showed patients mortality in ICU by Kaplan-Meier (KM) survival analysis.

## 3. Results

From January 1, 2019 to June 30, 2020 data were taken from 468 cases at Tibebe Ghion Specialized Teaching Hospital ICU registered log book through prepared questionnaire. We excluded 3(0.64%) cases with incomplete of outcome variable (patients’ death or safe discharged) data and 7(1.50%) cases with incomplete of independent variables (diagnosis at admission, need for mechanical ventilation, length of mechanical ventilation stay, length of stay in ICU) data and also 4 (0.87%) patients who died on arrival (within 2 hours of admission). Then 454 cases were entered in to ICU mortality analysis (Figure 1). Table 1 shows, socio-demographics and physical characteristics of the study participants. In this table, 36.3% of ICU admitted patients were served by mechanical ventilation. The pattern of patient admission and outcomes of adult intensive care unit were described in table 2. Among outline of these admissions, the head injury, respiratory problem, septic shock and stroke were constituted the higher ICU service mortality. Table 3 demonstrated the demographics and Clinical characteristics of logistic regression with a significant association of the infection, patients stayed in ICU and days on mechanical ventilation to ICU mortality. Figure 2 also showed the effects of ICU stay in days with the odds of mortality. In this figure, there was an increased probability of mortality when a patient waited more days in ICU. The number of days of patient stayed in intensive care unit and on mechanical ventilation was stated on figure 3. In this graph, both patients stay in ICU and days on mechanical ventilation demonstrated median (IQR) of 4(2-8) and 4(2-10) respectively.

**Table 1.**
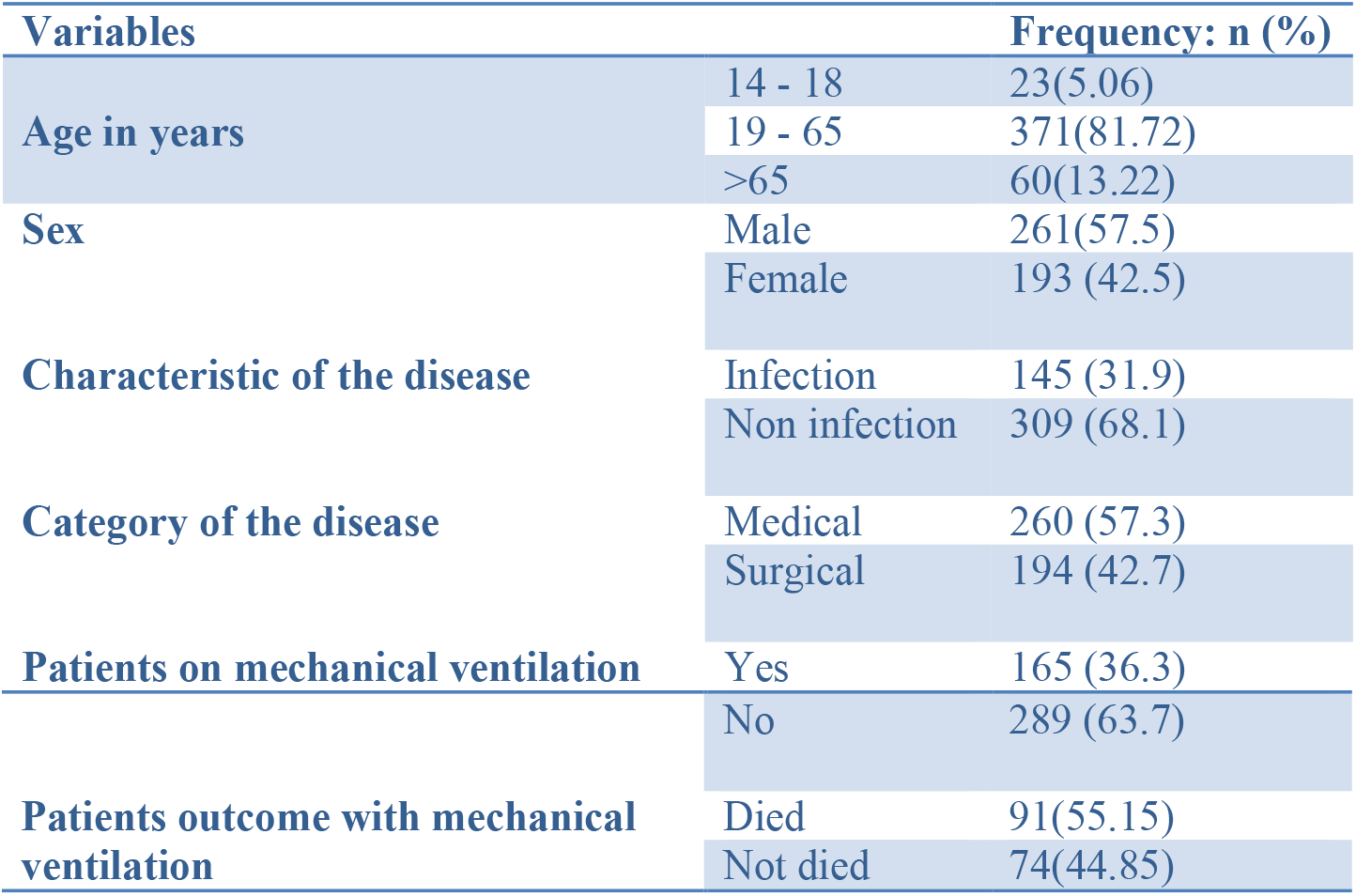
Socio-demographic and physical characteristics of study participants in intensive care unit in the period of January 1, 2019 – June 30, 2020.

**Table 2.** Pattern of admission and outcomes at discharge time in adult intensive care unit, Tibebe Ghion Specialized Teaching Hospital in the period of January 1, 2019 – June 30, 2020.

| Diagnosis | Outcome |  | Total (n) | Percentage of death over total sample size |
| --- | --- | --- | --- | --- |
|  | Died (n) | Not died (n) |  |  |
| Acute kidney injury | 4 | 4 | 8 | 0.88 |
| Cardiogenic shock | 2 | 4 | 6 | 0.44 |
| Congestive Heart Failure(CHF) | 3 | 18 | 21 | 0.66 |
| Liver problem(acute or chronic type and malignancy) | 7 | 15 | 22 | 1.54 |
| Diabetic mellitus (Type I and II) | 1 | 14 | 15 | 0.22 |
| Guillain Barre Syndrome (GBS) | 1 | 4 | 5 | 0.22 |
| Myocardial Infarction (MI) | 1 | 30 | 31 | 0.22 |
| Retro Viral Infection (RVI) | 10 | 8 | 18 | 2.20 |
| Post abdominal surgeries | 8 | 30 | 38 | 1.8 |
| Eclamsia | 2 | 1 | 3 | 0.44 |
| Pulmonary embolism | 0 | 7 | 7 | 0 |
| Epilepsy | 0 | 3 | 3 | 0 |
| Esophageal cancer | 2 | 4 | 6 | 0.44 |
| Fracture (upper/lower limb) | 1 | 5 | 6 | 0.22 |
| Head injury | 30 | 59 | 89 | 6.61 |
| Hemo/pneumothorax | 0 | 7 | 7 | 0 |
| Malaria | 1 | 4 | 5 | 0.22 |
| Meningitis | 2 | 2 | 4 | 0.44 |
| Pancreatic cancer | 1 | 3 | 4 | 0.22 |
| Peritonitis | 4 | 6 | 10 | 0.88 |
| Non-TB respiratory problems(Pneumonia/ARDS/Asthma) | 22 | 32 | 54 | 4.85 |
| Poison | 5 | 13 | 18 | 1.10 |
| Septic shock | 18 | 6 | 24 | 3.96 |
| Stroke | 10 | 9 | 19 | 2.20 |
| Tetanus | 3 | 10 | 13 | 0.66 |
| Post thyroid gland / parotid tumor surgeries | 2 | 9 | 11 | 0.44 |
| Tracheostomy | 2 | 5 | 7 | 0.44 |
| **Total** | 142 | 312 | 454 | 31.3% |

n=number of patients

**Table 3.**
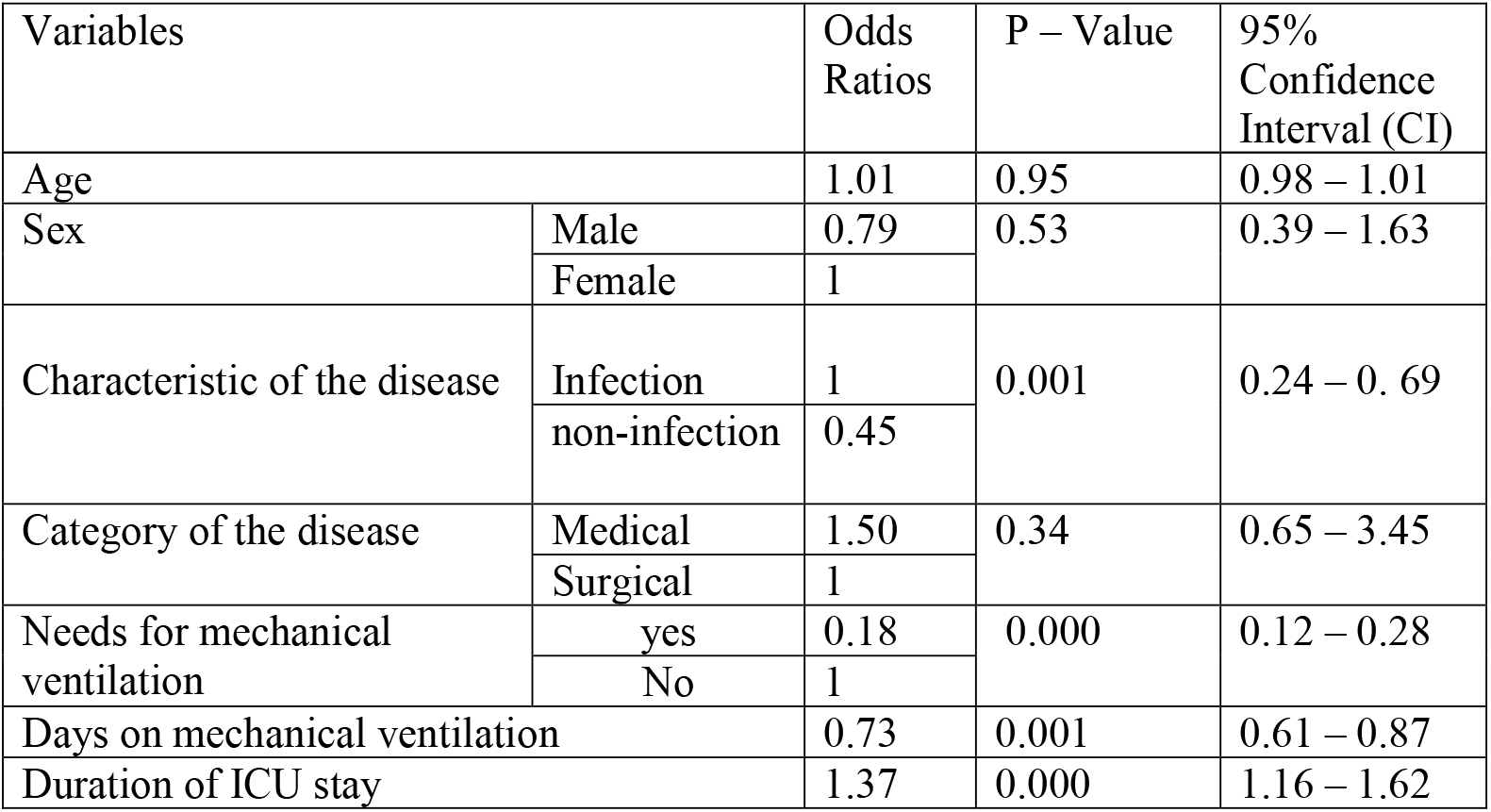
Demographics and Clinical characteristics of logistic regression in adult intensive care unit, Tibebe Ghion Specialized Teaching Hospital in the period of January 1, 2019 – June 30, 2020.

| Variables | Odds Ratios | P – Value | 95% Confidence Interval (CI) |
| --- | --- | --- | --- |

|  |  |  |  |  |
| --- | --- | --- | --- | --- |
| Age |  | 1.01 | 0.95 | 0.98 – 1.01 |
| Sex | Male | 0.79 | 0.53 | 0.39 – 1.63 |
|  | Female | 1 |  |  |
| Characteristic of the disease | Infection | 1 | 0.001 | 0.24 – 0.69 |
|  | non-infection | 0.45 |  |  |
| Category of the disease | Medical | 1.50 | 0.34 | 0.65 – 3.45 |
|  | Surgical | 1 |  |  |
| Needs for mechanical ventilation | yes | 0.18 | 0.000 | 0.12 – 0.28 |
|  | No | 1 |  |  |
| Days on mechanical ventilation |  | 0.73 | 0.001 | 0.61 – 0.87 |
| Duration of ICU stay |  | 1.37 | 0.000 | 1.16 – 1.62 |

**Figure 1.**
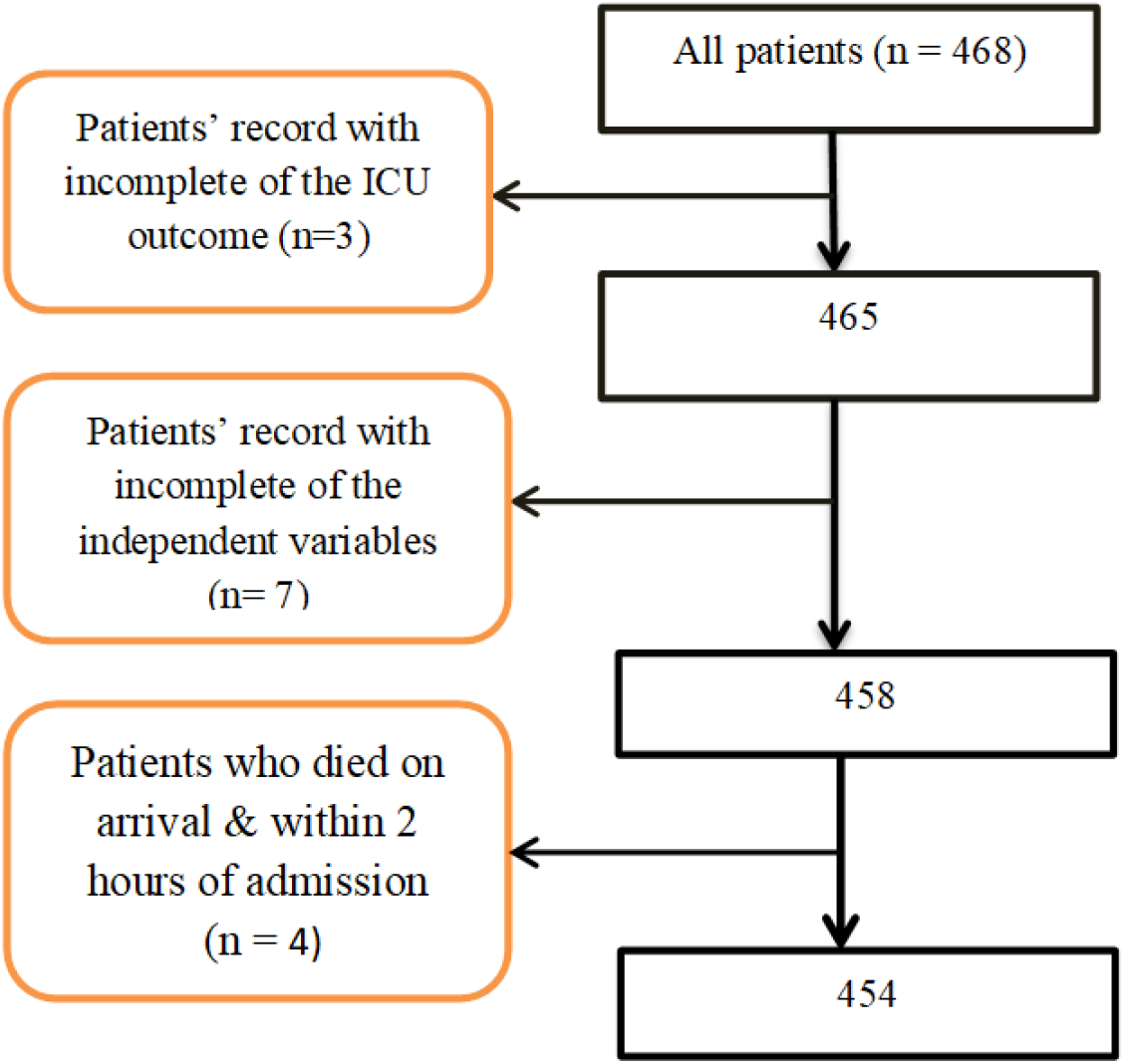
Number of cases with the missing of incomplete data

**Figure 2.**
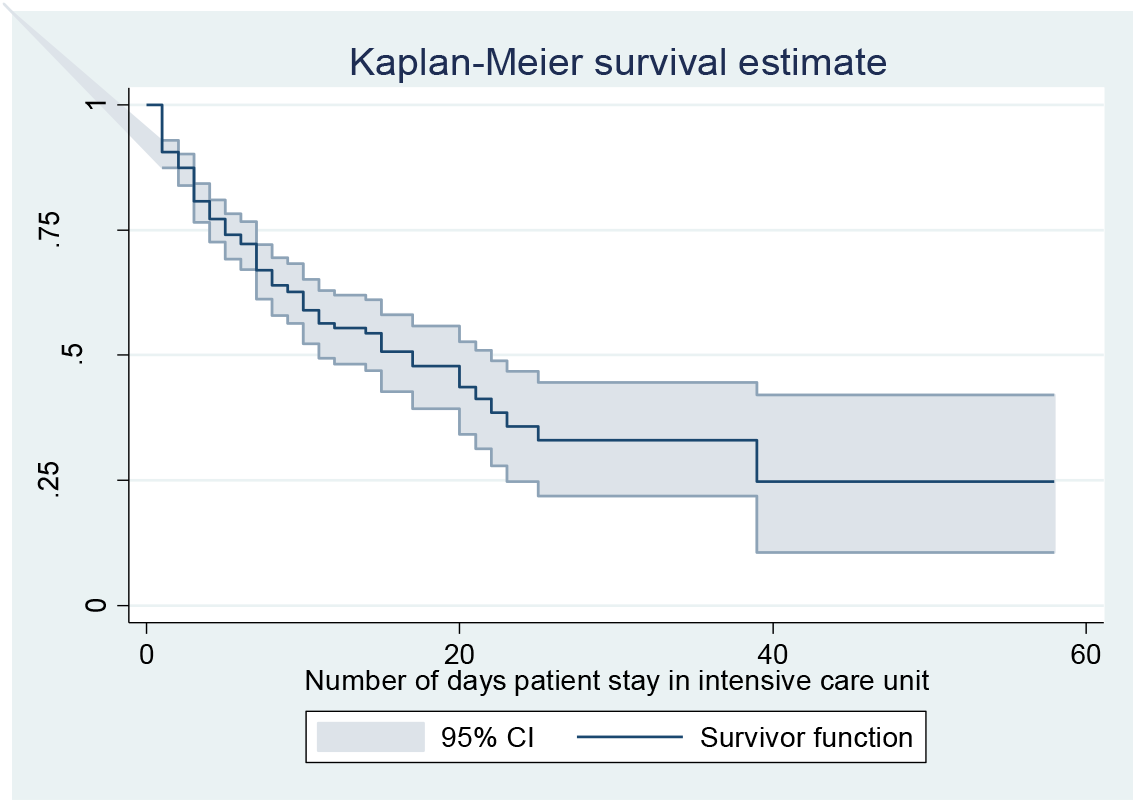
The Kaplan Meier survival estimate graph with the effect of patients’ ICU stay in days on odds of mortality

**Figure 3.**
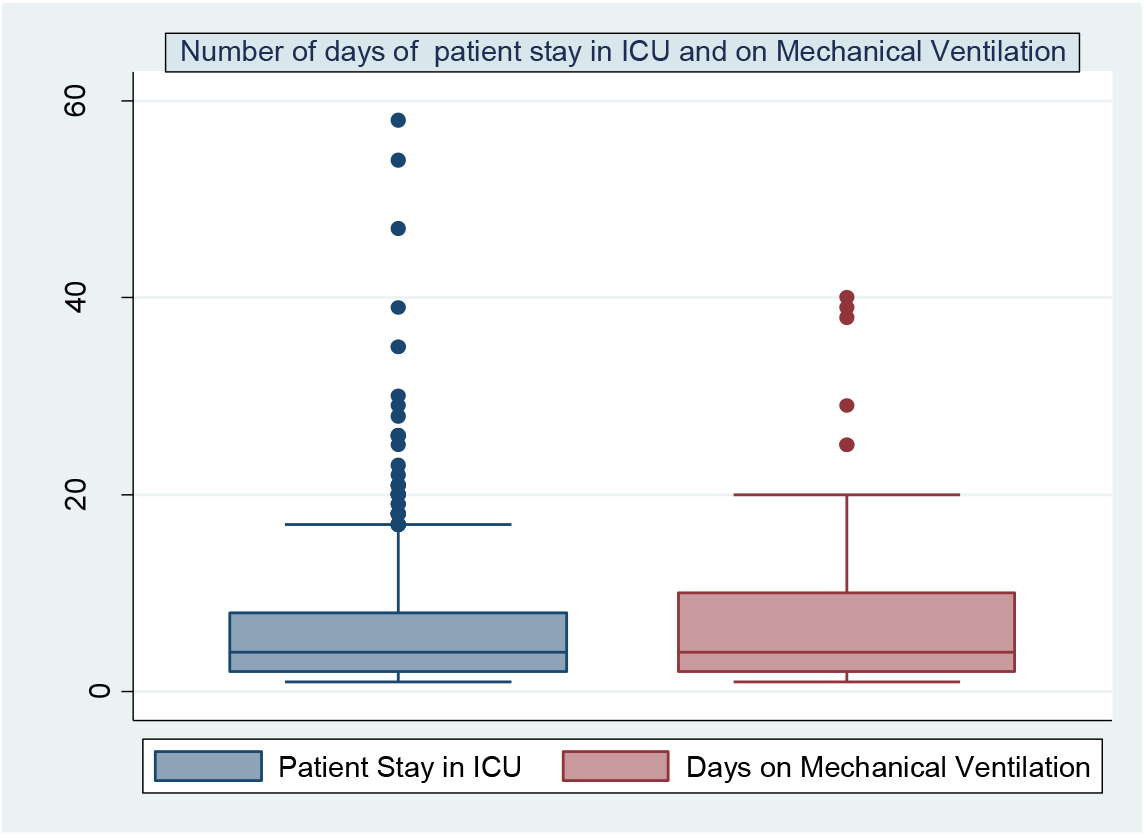
The Whisker Box plot graph of patients’ continued ICU care and being on mechanical ventilation, Tibebe Ghion Specialized Teaching Hospital in the period of January 1, 2019 – June 30, 2020

## 4. Discussion

The intention of the study was to identify the pattern and service outcome, and its predictors of patients admitted to adult ICU in Tibebe Ghion Specialized Teaching Hospital. In this study, the common leading causes of admission to ICU were head injury (19.6%) followed by non-tuberculosis respiratory problems (11.89%), post abdominal surgeries (8.37%) and myocardial infarction (6.82%). It was consistent to Tanzanian study on all age groups of admitted patients(14). The non-communicable diseases circumstance described for most of the ICU admissions, with trauma alone accounted that the 20% of non-communicable diseases burden was in sub-Saharan Africa countries(TanzaniaR35,36).However, it was not supported by an Ethiopian study conducted in Jimma University Hospital which showed that there was more number cardiovascular disorder cases than trauma and Post-surgical patients admitted in adult ICU(13). This discrepancy to our study could be related to the difference of surgical service setting up and expertise level.

There was a higher proportion (19.82%) of all deaths happened on head injury (6.61%), non-tuberculosis respiratory problems (Pneumonia/ARDS/Asthma) (4.85%), Septic shock (3.96%), retro viral infection (2.20%) and stroke (2.20%). This was……..

Furthermore, there was a significant association of patients stayed in ICU to the outcome variable (mortality) as a risk factor in our study with odds ratio (OR) = 1.37 [95% CI, 1.16 – 1.62]; *P* = 0.000).

The overall magnitude of ICU mortality (31.3%) was comparable to other studies of African countries (TanzaniaR30). Conversely, it was greatly higher than mortality stated in developed countries (TanzaniaR 28, 42).

The above figure 2 explained the effect of patient stayed in ICU on the log odds of ICU mortality rate compared to the earliest days’ time. There was appear to be much effect of mortality until 25 days of ICU stay but there was somewhat decline of ICU mortality between 25 and 40 days. This reduction of mortality rate could be linked with the lower number of patients continued in ICU care at a longer number of days.

Likewise, the median (IQR) length of in ICU stay (fig 3) was comparable to ICU studies of other Sub-Saharan Africa country (TanzaniaR30).

There were limitations to our study. First: there was documented data as good outcome in ICU registration log book for those patients rejected the mechanical ventilation. Second: some other patients left the ICU care by refusal and recorded as not died. These phenomena could affect the outcome of ICU (mortality) and to be considered as a confounding variable.

## 5. Conclusion

The mortality rate of our adult ICU was 31.3% with the most common causes of admission and deaths being on head injury. The highly statistical significant predictors of ICU mortality were infection, needs for mechanical ventilation, ICU stay and days on mechanical ventilation. The hospital should emphasis on improvement plans of ICU care on the higher mortality recorded cases of head injury, non TB respiratory problems and septic shock.

## Data Availability

All data are available from the corresponding author upon reasonable request.

## Abbreviations

CI: Confidence Interval,
ERB: Ethical Review Board,
ICU: Intensive Care Unit,
IQR: Inter Quartile Range,
OR: Odds Ratio,
SD: Standard Deviation,
TB: Tuberculosis

## Acknowledgments

We want to thank anesthetists and anesthesia students working at Tibebe Ghion Specialized Hospital for aiding the data collection.

## Author contribution

**AS Endeshaw**: Took part in conceptualization, methodology, formal analysis, investigation, resources, data curation, writing - original manuscript draft, writing – review & editing, visualization, and supervision. **FT Kume and MT Molla**: Took part in methodology, formal analysis, investigation, writing review & editing, and visualization. **GA Zeru**: Manuscript writing, paper revision, editing.

## Data sharing statement

Data are available from the corresponding author upon reasonable request.

## Ethical Approval

The ethical approval of this research, including an informed consent waiver, was given by Bahir Dar University, College of Medicine and Health Sciences institutional review board (IRB).

## References

1. Berthelsen P, Cronqvist M. The first intensive care unit in the world: Copenhagen 1953. Acta Anaesthesiologica Scandinavica. 2003;47(10):1190–5.

2. Rao SM, Suhasini T. Organization of intensive care unit and predicting outcome of critical illness. Indian journal of anaesthesia. 2003;47(05):328–37.

3. Dunser MW BI, Ganbold L. A review and analysis of intensive care medicine in the least developed countries. Crit Care Med. 2006;34 1234–42.

4. Riviello ED LS, Achieng L, Newton MW. Critical care in resource-poor settings: Lessons learned and future directions. Crit Care Med. 2011.

5. Towey R, Ojara S. Intensive care in the developing world. Anaesthesia. 2007;62:32–7.

6. Team WER. Ebola virus disease in West Africa—the first 9 months of the epidemic and forward projections. New England Journal of Medicine. 2014;371(16):1481–95.

7. Murray CJ. Phil., Alan D. Lopez. Measuring the Global Burden of Disease. New England Journal of Medicine. 2013;369:448–57.

8. Annez PC, Linn JF. An agenda for research on urbanization in developing countries: a summary of findings from a scoping exercise: The World Bank; 2010.

9. Baelani I, Jochberger S, Laimer T, Otieno D, Kabutu J, Wilson I, et al. Availability of critical care resources to treat patients with severe sepsis or septic shock in Africa: a self-reported, continent-wide survey of anaesthesia providers. Critical care. 2011;15(1):R10.

10. Dünser MW, Baelani I, Ganbold L. A review and analysis of intensive care medicine in the least developed countries. Critical care medicine. 2006;34(4):1234–42.

11. Smith Z, Ayele Y, McDonald P. Outcomes in critical care delivery at Jimma University Specialised Hospital, Ethiopia. Anaesthesia and intensive care. 2013;41(3):363–8.

12. Seman Kedir AB, Tola Bayisa, Tewodros Wuletaw Admission patterns and outcomes in the medical intensive care unit of st. paul’s hospital millennium medical college, Addis ababa, Ethiopia. Ethiop Med J,. 2017(1):19–26.

13. Asrat Agalu MW, Yemane Ayele, Worku Bedada. Reasons for admission and mortalities following admissions in the intensive care unit of a specialized hospital, in Ethiopia. Int J Med Med Sci. 2014;6(9):195–200.

14. Sawe HR, Mfinanga JA, Lidenge SJ, Mpondo BC, Msangi S, Lugazia E, et al. Disease patterns and clinical outcomes of patients admitted in intensive care units of tertiary referral hospitals of Tanzania. BMC international health and human rights. 2014;14(1):26.

15. Adudu O, Ogunrin O, Adudu O. Morbidity and mortality patterns among neurological patients in the intensive care unit of a tertiary health facility. Annals of African Medicine. 2007;6(4).

